# Insights from Single-Cell RNA-seq: Identifying the Actin Gene Family as Novel Drivers in Parkinson’s Disease

**DOI:** 10.1101/2024.02.12.24302696

**Authors:** Saed Sayad, Mark Hiatt, Hazem Mustafa

## Abstract

**Background:** Parkinson’s disease (PD) is the second most common neurodegenerative disorder, affecting millions of individuals worldwide. The complex etiology of PD involves a combination of genetic and environmental factors. The Actin family encompasses a group of highly conserved cytoskeletal proteins that play a crucial role in maintaining cellular structure and function. Actin proteins are involved in various cellular processes, including cell motility, vesicle trafficking, and synaptic transmission. This paper delves into the exploration of the Actin family of genes, revealing their potential as key contributors to Parkinson’s Disease through the application of single-cell RNA-seq.

**Method:** We obtained single-cell transcriptomes (*GSE237133*) from the NIH portal website. We conducted an extensive comparative analysis of single-cell transcriptomes derived from Parkinson’s disease organoids and two control organoids to identify differentially expressed genes, pathways, and gene ontology terms.

**Results:** We conducted a comparative analysis of single-cell transcriptomes from Parkinson’s disease organoid and two control organoids, aiming to identify differentially expressed genes, pathways, and gene ontology items. In comparing the PD organoid with the control organoid, we observed that the ACTB and ACTG1 genes were common among 18 of the top 20 upregulated KEGG pathways and among 15 of the top 20 upregulated Reactome pathways. Additionally, when comparing the PD organoid with the isogenic control organoid, we found the ACTB and ACTG1 genes shared among 19 out of the top 20 pathways and among 19 out of the top 20 upregulated Reactome pathways. An additional noteworthy finding includes the overexpression of several “Mitochondrially Encoded NADH” family genes in the PD organoid cells compare to the control organoids cells.

**Conclusion:** The Actin family of genes in general and ACTB and ACTG1 genes in particular emerges as a potential new player in the convoluted landscape of Parkinson’s disease. Further research is needed to elucidate the precise mechanisms through which Actin dysregulation contributes to PD pathology and to develop targeted therapeutic approaches. Unraveling the connections between Actin and PD may pave the way for innovative strategies to intervene in the disease process, ultimately improving the lives of individuals affected by Parkinson’s disease.

## Introduction

Parkinson’s disease (PD) stands as the second most prevalent neurodegenerative disorder globally, impacting millions with its multifaceted etiology influenced by both genetic and environmental factors. The Actin family encompasses a group of highly conserved cytoskeletal proteins that play a crucial role in maintaining cellular structure and function. Actin proteins are involved in various cellular processes, including cell motility, vesicle trafficking, and synaptic transmission (1). Several lines of evidence point to the dysregulation of Actin dynamics in the brains of individuals with PD and revealed altered Actin expression and distribution in the substantia nigra, a key region affected in PD (2). Additionally, genetic studies have identified variations in Actin family genes in PD patients, highlighting their potential role as genetic drivers of the disease. Dysregulation of Actin polymerization and depolymerization processes may contribute to the formation of Lewy bodies and the degeneration of dopaminergic neurons (3). Understanding the role of the Actin family of genes in PD opens avenues for developing targeted therapeutic interventions. Modulating Actin dynamics through small molecules or gene therapies may provide novel strategies to slow or halt the progression of PD. Additionally, identifying Actin-related biomarkers could facilitate early diagnosis and intervention, improving outcomes for individuals at risk of developing PD. This paper unveils the potential contribution of the Actin family to Parkinson’s disease pathology using single-cell RNA-seq data.

## Data

We downloaded single-cell RNA-seq (*GSE237133*) from the NIH portal website. It encompassed single cell profile of human iPSC-derived midbrain organoids from healthy individual (control), Parkinson disease patient harboring a Miro1R272Q mutation and its respective isogenic control (**Figure 1**).

**Figure 1:**
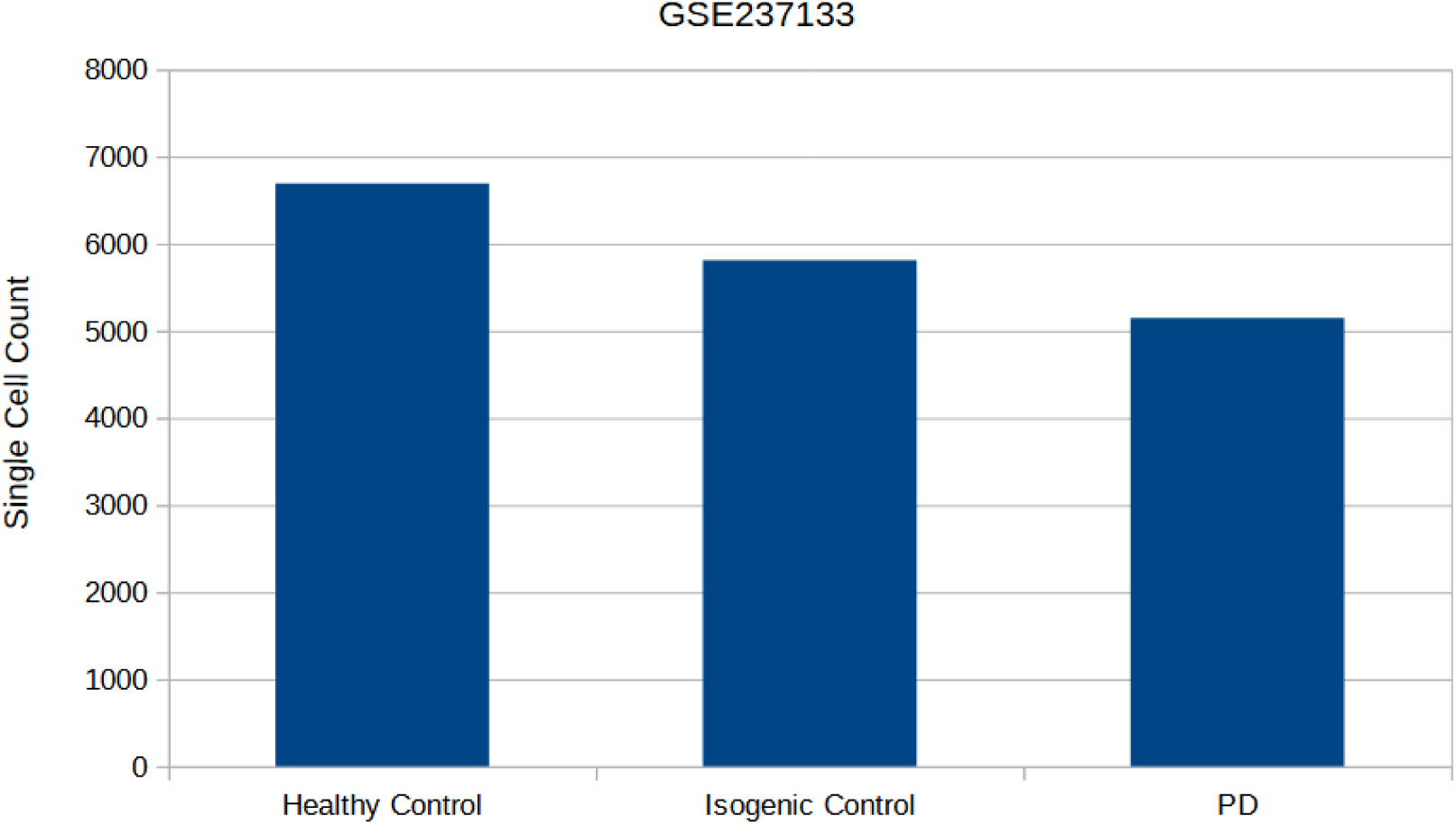
Healthy control, isogenic control and Parkinson’s disease (PD) single cells count (4).

## Data Analysis

In this study, we performed a comprehensive comparative analysis of single-cell transcriptomes obtained from Parkinson’s Disease (PD) organoids and two control organoids. Our central aim revolved around the identification of differentially expressed genes, pathways, and gene ontology items. Through this thorough analysis, we sought to illuminate potential molecular intricacies that could contribute to a deeper understanding of the underlying mechanisms in Parkinson’s Disease.

## Healthy Control single cells against PD single cells

Using the t-test, we compared the healthy control data (6693 single cells from organoid) with the Parkinson’s disease data (5148 single cells from organoid) to find differentially expressed genes, pathways and gene ontology items. Out of many differentially expressed genes, we selected top ten upregulated and downregulated genes (**Table 1**) that represent a diverse range of biological processes and functions but are especially connected to the immune system.

**Table 1:**
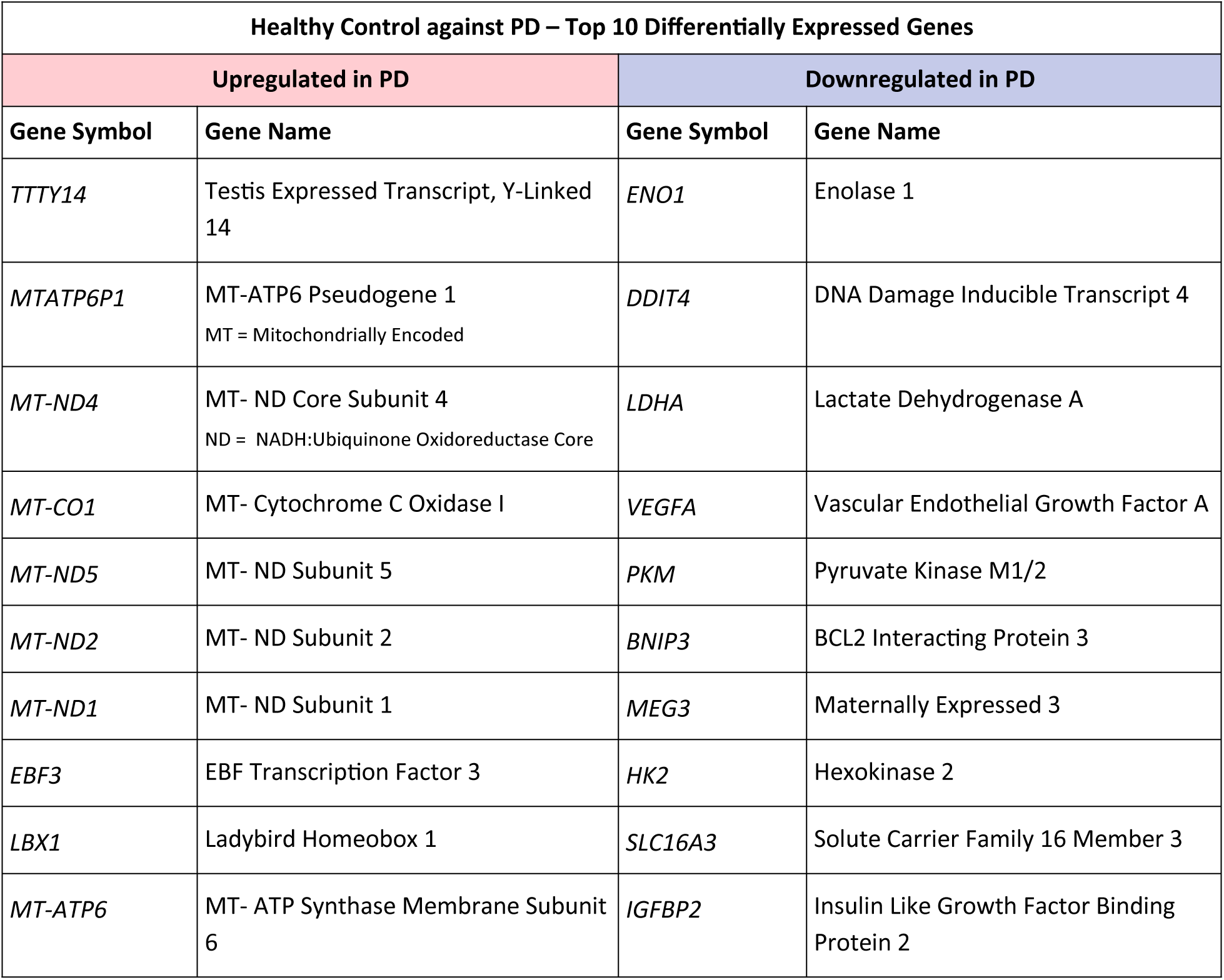
Top ten upregulated and downregulated genes through Healthy Control and PD gene expression analysis.

Regarding the upregulated genes, TTTY14 has been reported as one of the top five most overexpressed genes within dopaminergic neurons (5). Multiple studies consistently highlight the significant involvement of mitochondria and mitochondrial dysfunction in the development of Parkinson’s disease (6). Notably, research has demonstrated that the overexpression of Transcription factor EB (EBF3) effectively hinders neurodegeneration in experimental synucleinopathies (7). Moreover, the ladybird homeobox (LBX1) genes are essential for the specification of a subpopulation of neural cells (8).

The downregulation of certain genes, particularly those associated with elevated neuron-specific enolase (NSE), has been linked to detrimental effects on the extracellular matrix (ECM), inflammatory glial cell proliferation, and actin remodeling. These processes impact the migration of activated macrophages and microglia to the injury site, ultimately promoting neuronal cell death (9). DNA damage is also implicated in Parkinson’s disease, contributing to protein aggregation, senescence, cell death, and neuroinflammation (10). A hallmark of aging is the elevation of brain lactate, attributed to a shift in the lactate dehydrogenase A/B ratio (11). Notably, various experiments suggest a potential neuroprotective role for vascular endothelial growth factor (VEGF) in Parkinson’s disease treatment (12). Mutations in the Parkin gene, a ubiquitin E3 ligase, are prevalent in autosomal recessive early-onset Parkinson’s disease. Parkin’s unique substrate, pyruvate kinase M2 (PKM2), has been identified through biochemical purification, revealing its role in regulating the glycolysis pathway and influencing cell metabolism— providing new avenues for PD therapy (13). The BNIP3 gene encodes a mitochondrial protein acting as a pro-apoptotic factor with implications for both natural and pathological neuronal death (14). Long non-coding RNA MEG3, expressed at lower levels in the plasma of PD patients, is closely linked to the severity of non-motor symptoms, cognitive decline, and PD stage (15). Upregulated hexokinase 2 expression has been associated with the apoptosis of dopaminergic neurons in Parkinson’s disease, driven by increased lactate production (16). Additionally, the SLC2A13 gene has been identified as a risk factor for PD in the Chinese population (17). An Experiment using insulin-like growth factor-2 has shown promise in restoring dopamine neuron markers, reducing α-synuclein aggregation, and alleviating insulin-like growth factor-2 receptor downregulation in PD cell models and mouse models induced by 6-hydroxydopamine (18).

**Table 2** illustrates the prominent KEGG pathways that are both upregulated and downregulated in PD organoids as opposed to Healthy Control. All of the top upregulated pathways show a direct connection to PD (click on the pathway to see the reference). Significantly, the ACTB and ACTG1 genes are found to be shared among 9 out of the top 10 upregulated KEGG pathways and 18 out of the top 20. Thorne et al. have detailed the association between Miro, alpha-Synuclein, and Actin in their study (3), adding depth to our understanding of these findings. The majority of the top downregulated KEGG pathways show a direct connection to PD (click on the pathway to see the reference). Notably, the Hexokinases (HK) and HLAs genes shared among majority of the top 10 downregulated KEGG pathways. Hexokinase 2 (HK2) is on the list of the top 10 downregulated genes (Table 1). Regarding HLAs genes, considering the chronic inflammatory characteristics of PD, humoral immunity may significantly influence its progression, while T cell immunity might play a more crucial role in disease onset (19).

**Table 2:** Top ten upregulated and downregulated KEGG pathways in Healthy Control compared to PD single cells.

| Healthy Control against PD – Top 10 Differentially Expressed KEGG Pathways |  |
| --- | --- |
| Upregulated in PD | Downregulated in PD |
| Arrhythmogenic right ventricular cardiomyopathy | Fructose and mannose metabolism |
| Viral myocarditis | Neomycin, kanamycin and gentamicin biosynthesis |
| Hypertrophic cardiomyopathy | Glycolysis / Gluconeogenesis |
| African trypanosomiasis | Central carbon metabolism in cancer |
| Platelet activation | Maturity onset diabetes of the young |
| Dilated cardiomyopathy | HIF-1 signaling pathway |
| Adherens junction | Allograft rejection |
| Hippo signaling pathway | Graft-versus-host disease |
| Thyroid hormone signaling pathway | Autoimmune thyroid disease |
| Bacterial invasion of epithelial cells | Taurine and hypotaurine metabolism |

**Table 3** illustrates the prominent Reactome pathways that are both upregulated and downregulated in PD organoids as opposed to Healthy Control. The majority of the top upregulated pathways show a direct connection to PD (click on the pathway to see the reference). Notably, like KEGG pathways, the ACTB and ACTG1 genes are found to be shared among 8 out of the top 10 upregulated Reactome pathways and 15 out of the top 20. Moreover, the majority of the top downregulated Reactome pathways show a direct connection to PD as well (click on the pathway to see the reference) and VEGFs and SLCs genes shared among majority of the top 10 downregulated pathways. Both VEGFA and SLC16A3 genes are on the list of the top 10 downregulated genes (Table 1).

**Table 3:** Top ten upregulated and downregulated Reactome pathways for Healthy Control against PD single cells.

| Healthy Control against PD – Top 10 Differentially Expressed Reactome Pathways |  |
| --- | --- |
| Upregulated in PD | Downregulated in PD |
| MECP2 regulates transcription factors | Manipulation of host energy metabolism |
| Interaction between L1 and Ankyrins | Cellular hexose transport |
| Receptor-type tyrosine-protein phosphatases | VEGF ligand-receptor interactions |
| Cell-extracellular matrix interactions | VEGF binds to VEGFR leading to receptor dimerization |
| Adherens junctions interactions | GABA synthesis |
| Cell-cell junction organization | Lactose synthesis |
| Formation of annular gap junctions | Defective SLC2A1 causes GLUT1 deficiency syndrome 1 (GLUT1DS1) |
| Gap junction degradation | Regulation of gene expression in early pancreatic precursor cells |
| Signaling by high-kinase activity BRAF mutants | Vitamin C (ascorbate) metabolism |
| EPH-ephrin mediated repulsion of cells | MECP2 regulates transcription of genes involved in GABA signaling |

On the list of top 10 upregulated gene ontology terms (**Table 4**), there is a notable upregulation in biological processes (BP) related to synaptic processes. This includes positive regulation of synapse assembly, synaptic membrane adhesion, and positive regulation of synapse maturation. These changes suggest a significant impact on synaptic function and organization. Understanding these alterations in gene expression could provide valuable insights into the synaptic dysfunction associated with PD. Meanwhile, VEGFs and ENOLs genes shared among majority of the top 10 downregulated biological processes (BP), molecular function (MF) and cellular components (CC). Both VEGFA and ENOL1 genes are notably part of the list of the top 10 downregulated genes (Table 1).

**Table 4:** Top ten upregulated and downregulated Gene Ontology terms for Healthy Control against PD single cells.

| Healthy Control against PD – Top 10 Differentially Expressed Gene Ontology terms |  |
| --- | --- |
| Upregulated in PD | Downregulated in PD |
| (BP) regulation of postsynaptic density assembly | (BP) positive regulation of muscle contraction |
| (BP) regulation of transcription from RNA polymerase II promoter involved in spinal cord association | (BP) positive regulation of plasminogen activation |
| (BP) positive regulation of synapse assembly | (MF) phosphopyruvate hydratase activity |
| (BP) synaptic membrane adhesion | (CC) phosphopyruvate hydratase complex |
| (BP) development of primary female sexual characteristics | (BP) negative regulation of intracellular signal transduction |
| (BP) neuron cell-cell adhesion | (BP) post-embryonic animal organ development |
| (BP) positive regulation of synapse maturation | (BP) primitive erythrocyte differentiation |
| (BP) negative regulation of cell proliferation involved in contact inhibition | (BP) positive regulation of blood vessel endothelial cell proliferation involved in sprouting angiog |
| (BP) regulation of nuclear cell cycle DNA replication | (BP) negative regulation of establishment of endothelial barrier |
| (BP) regulation of presynapse assembly | (BP) positive regulation of protein kinase D signaling |

## Isogenic Control single cells against PD single cells

We employed the t-test to analyze the isogenic control data (5811 single cells from organoid) in comparison to the Parkinson’s disease data (5148 single cells from organoid). Our aim was to identify genes, pathways, and gene ontology items that exhibit differential expressions. From the pool of differentially expressed genes, we curated a list of the top ten upregulated and downregulated genes (**Table 5**). The genes highlighted in bold, such as TTTY14 and VEGFA, exhibit commonality with the genes listed in Table 1.

**Table 5:**
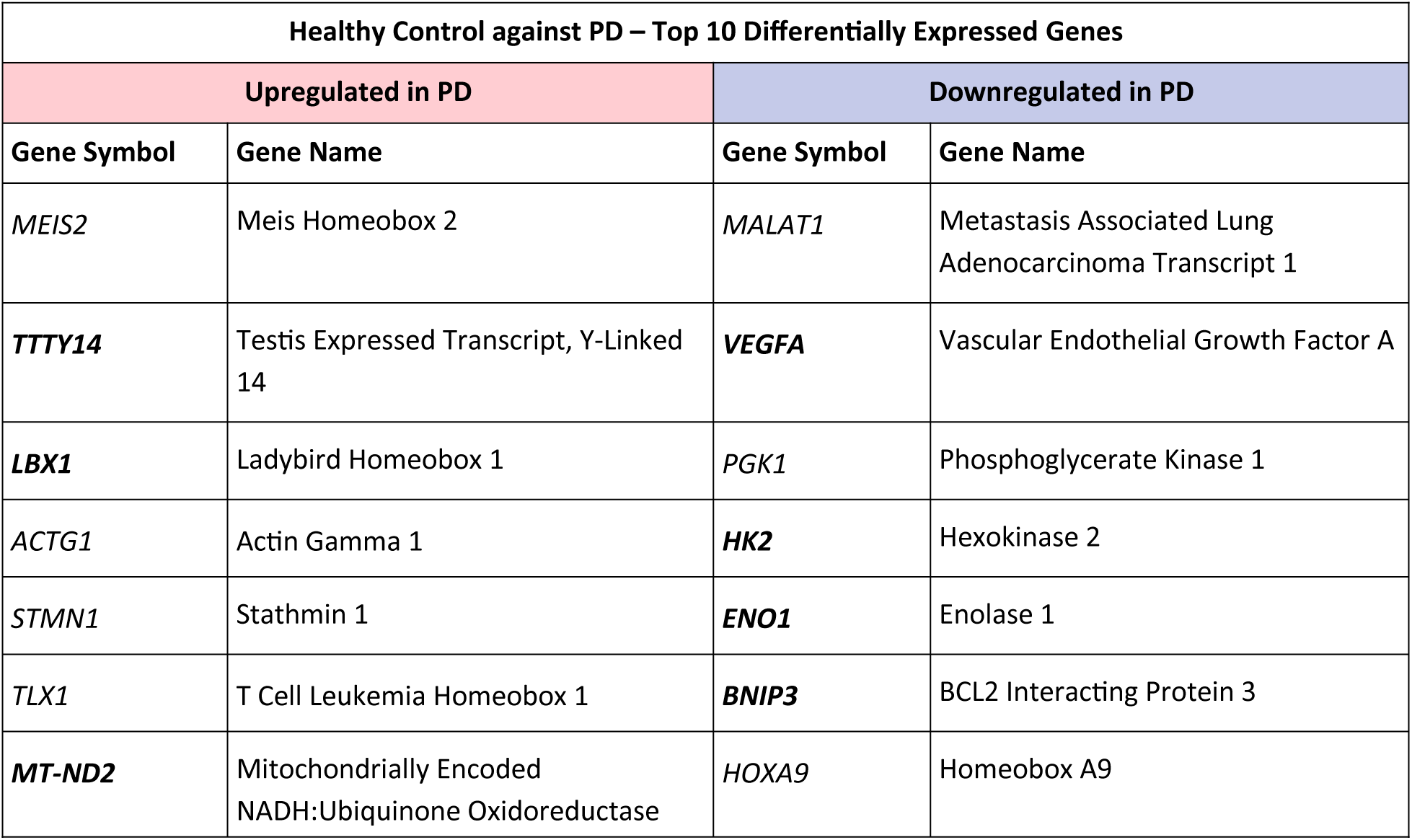
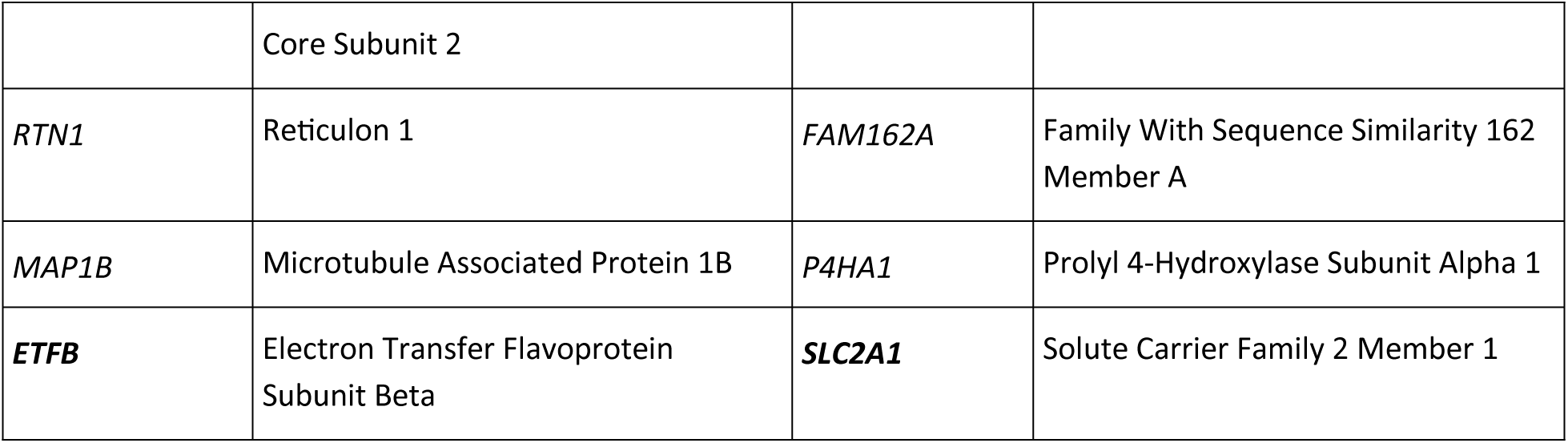
Top ten upregulated and downregulated genes through Isogenic Control and PD gene-expression analysis.

As for the upregulated genes, MEIS1 dysfunction contributes to motor circuit abnormalities, and the effectiveness of D2-like agonists in treating restless legs syndrome may involve influencing various dopaminergic receptors or normalizing downstream defects in the motor circuits (20). ACTG1 is a key actin gene in this study involved in the establishment of the cytoskeleton and autophagy pathway in PD (21). Proteins of the stathmin family affect microtubule dynamics regulating the assembly and the dismantling of tubulin in Motor neuron diseases (22). It has been suggested RTN-1C protein as a promising target in the treatment of different pathologies such as cancer or neurodegenerative disorders (23).

Regarding the downregulated gene, MDMA promotes MALAT1 expression and inhibits the targeted downregulation of MEKK3 by miR-124, resulting in upregulation of the expression of MEKK3 and finally jointly promoting PD progression (24). In PD induced neuronal metabolic deficits, phosphoglycerate kinase (PGK1) serves as a crucial focal point (25). Moreover, it has been shown that inhibition of prolyl hydroxylase is a promising therapeutic strategy for PD through improving the mitochondrial function under oxidative stress (26).

In **Table 6**, we spotlight the key KEGG pathways both upregulated and downregulated in PD when compared to the Isogenic Control single cells. Notably, a substantial proportion of these pathways align with the top 10 pathways identified in Healthy Control versus PD. Interestingly, the ACTB and ACTG1 genes are found to be shared among 10 out of the top 10 upregulated KEGG pathways and 19 out of the top 20. Similar to the Healthy Control versus PD scenario, the Hexokinases (HK) and HLAs genes are commonly shared among the majority of the top 10 downregulated KEGG pathways.

**Table 6:** Top ten upregulated and downregulated KEGG pathways for Isogenic Control against PD single cells.

| Isogenic Control against PD – Top 10 Differentially Expressed KEGG Pathways |  |
| --- | --- |
| Upregulated in PD | Downregulated in PD |
| **Arrhythmogenic right ventricular cardiomyopathy** | **Neomycin, kanamycin and gentamicin biosynthesis** |
| **Hypertrophic cardiomyopathy** | **Allograft rejection** |
| **Viral myocarditis** | **Autoimmune thyroid disease** |
| Pathogenic Escherichia coli infection | **Graft-versus-host disease** |
| **Bacterial invasion of epithelial cells** | **Taurine and hypotaurine metabolism** |
| Tight junction | **HIF-1 signaling pathway** |
| Shigellosis | **Glycolysis / Gluconeogenesis** |
| **Adherens junction** | **Fructose and mannose metabolism** |
| Apoptosis | **Central carbon metabolism in cancer** |
| **Platelet activation** | Intestinal immune network for IgA production |

In **Table 7**, we show the top 10 Reactome pathways, showcasing both upregulated and downregulated patterns in Parkinson’s Disease (PD) compared to the Isogenic Control single cells. Mirroring the findings in KEGG pathways, a significant portion of these pathways aligns with those identified in the Healthy Control versus PD analysis. Notably, similar to the KEGG pathways, ACTB and ACTG1 genes are consistently present in 10 out of the top 10 upregulated Reactome pathways and 19 out of the top 20. Furthermore, mirroring the Healthy Control versus PD comparison, VEGFs and SLCs genes are recurrently shared among the majority of the top 10 downregulated Reactome pathways.

**Table 7:** Top ten upregulated and downregulated Reactome pathways for Isogenic Control against PD single cells.

| Isogenic Control against PD – Top 10 Differentially Expressed Reactome Pathways |  |
| --- | --- |
| Upregulated in PD | Downregulated in PD |
| Nuclear signaling by ERBB4 | Manipulation of host energy metabolism |
| Cell-extracellular matrix interactions | Cellular hexose transport |
| Interaction between L1 and Ankyrins | Defective SLC2A1 causes GLUT1 deficiency syndrome 1 (GLUT1DS1) |
| Formation of annular gap junctions | Lactose synthesis |
| Adherens junctions interactions | VEGF ligand-receptor interactions |
| Gap junction degradation | VEGF binds to VEGFR leading to receptor dimerization |
| Cell-cell junction organization | GABA synthesis |
| EPHB-mediated forward signaling | Vitamin C (ascorbate) metabolism |
| EPH-ephrin mediated repulsion of cells | MECP2 regulates transcription of genes involved in GABA signaling |
| Signaling by high-kinase activity BRAF mutants | VLDL assembly |

The top 10 upregulated and downregulated Gene Ontology terms are presented in **Table 8**. Notably, only two genes from this table coincide with those in Table 4. However, when comparing the top 100 Gene Ontology terms between Healthy Control and PD, as well as Isogenic Control and PD, we noted a substantial overlap of over 70%. A noteworthy finding is that the ACTG1 gene is present in 5 out of the top 10 upregulated Gene Ontology terms, while VEGFA is associated with 8 out of the top 10 Gene Ontology terms.

**Table 8:**
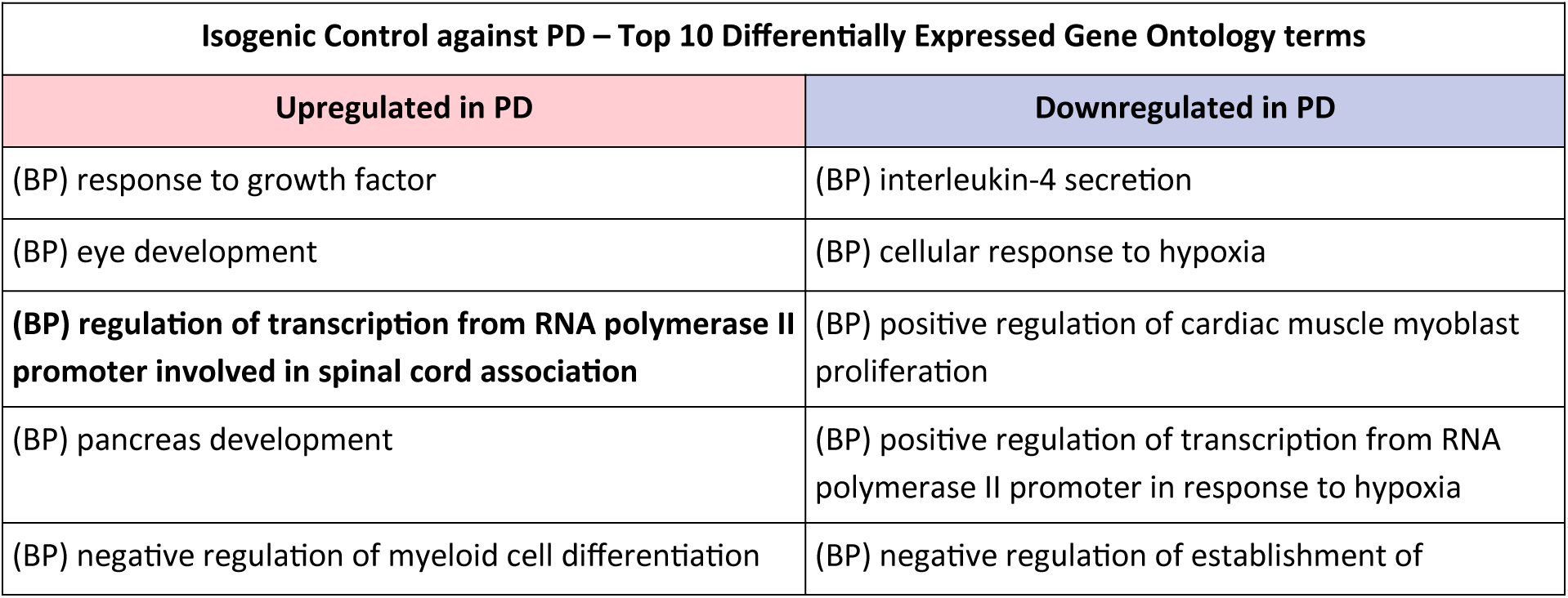
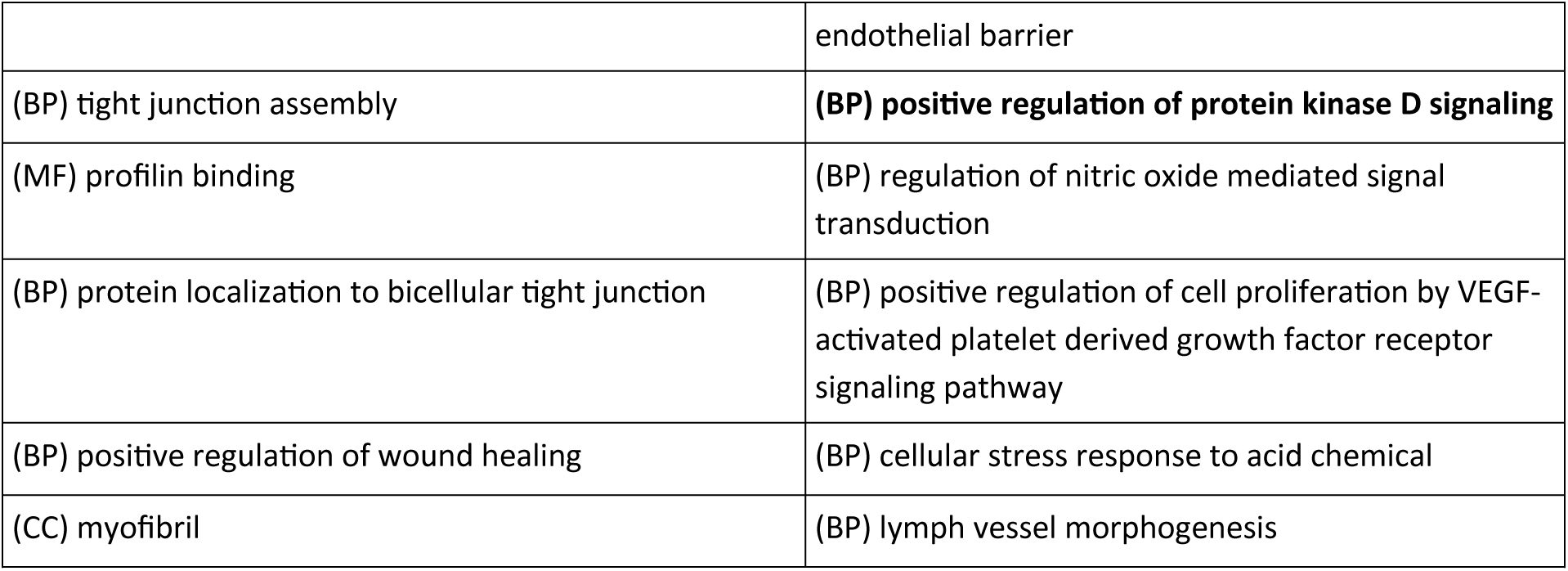
Top ten upregulated and downregulated Gene Ontology terms for Isogenic Control against PD single cells.

## Discussion

The comprehensive analysis of single-cell RNA-seq data from Parkinson’s Disease (PD) and controls organoids sheds light on the intricate molecular landscape underlying the disease. The identification of differentially expressed genes, pathways, and gene ontology terms provides valuable insights into potential mechanisms and therapeutic targets.

The upregulated genes in Parkinson’s Disease, emphasize the involvement of mitochondrial dysfunction, neuroprotection, and neural cell specification in PD pathology. On the other hand, downregulated genes like NSE-associated genes and BNIP3 highlight links to extracellular matrix effects, inflammation, and apoptosis. These findings align with existing literature on PD, reinforcing the multifaceted nature of the disease. The KEGG pathway analysis further strengthens the connection between Actin dynamics and PD, with ACTB and ACTG1 genes consistently shared among the top upregulated pathways. Thorne et al.’s study on the association between Miro, alpha-Synuclein, and Actin adds depth to these observations. The downregulated KEGG pathways, particularly those involving Hexokinases and HLAs genes, emphasize the impact of immune response and inflammatory characteristics in PD progression. The Reactome pathway analysis echoes the KEGG findings, emphasizing the significance of Actin-related genes in PD. Notably, the involvement of VEGFs and SLCs genes in downregulated Reactome pathways hints at the potential role of vascular and metabolic factors in PD pathogenesis. The Gene Ontology terms highlight the impact on synaptic processes in upregulated pathways, providing insights into synaptic dysfunction in PD. Meanwhile, the downregulated terms, including VEGFs and ENOLs genes, suggest broader implications for biological processes, molecular functions, and cellular components.

The comparison between PD and isogenic controls reveals commonalities in the upregulated genes, pathways, and gene ontology terms. This consistency underscores the robustness of the findings and emphasizes the potential relevance of these genes in PD pathogenesis.

## Summary

This study delves into the molecular landscape of Parkinson’s disease using single-cell RNA-seq data, highlighting the pivotal role played by the Actin family genes, including ACTB and ACTG1. The identified genes, pathways, and ontology terms not only deepen our understanding of PD mechanisms but also open avenues for targeted therapeutic interventions and potential early diagnostic biomarkers. Further research is imperative to unveil the precise mechanisms through which Actin dysregulation contributes to Parkinson’s disease pathology and to formulate specific therapeutic approaches. By unraveling the connections between Actin and Parkinson’s disease, this research could pave the way for innovative strategies to intervene in the disease process, ultimately enhancing the quality of life for individuals affected by Parkinson’s disease.

## Data Availability

All data produced are available online at:
https://www.ncbi.nlm.nih.gov/geo/query/acc.cgi?acc=GSE237133

## References

1. Murphy AC, Young PW. The actinin family of actin cross-linking proteins - a genetic perspective. Cell Biosci. 2015 Aug 25;5:49. Doi: 10.1186/s13578-015-0029-7. Oliveira da Silva MI, Liz MA. Linking Alpha-Synuclein to the Actin Cytoskeleton: Consequences to Neuronal Function. Front Cell Dev Biol. 2020 Aug 12;8:787.

2. Walker DG, Lue LF, Serrano G, Adler CH, Caviness JN, Sue LI, Beach TG. Altered Expression Patterns of Inflammation-Associated and Trophic Molecules in Substantia Nigra and Striatum Brain Samples from Parkinson’s Disease, Incidental Lewy Body Disease and Normal Control Cases. Front Neurosci. 2016 Jan 14;9:507.

3. Thorne NJ, Tumbarello DA. The relationship of alpha-synuclein to mitochondrial dynamics and quality control. Front Mol Neurosci. 2022 Aug 26;15:947191.

4. Axel Chemla, Giuseppe Arena, Ginevra Sacripanti, Kyriaki Barmpa, Alise Zagare, Pierre Garcia, Paul Antony, Jochen Ohnmacht, Alexandre Baron, Jaqueline Jung, Anne-Marie Marzesco, Manuel Buttini, Thorsten Schmidt, Anne Grünewald, Jens C. Schwamborn, Rejko Krüger, Claudia Saraiva. Parkinson’s disease-related Miro1 mutation induces mitochondrial dysfunction and loss of dopaminergic neurons in vitro and in vivo. BioRxiv 2023.12.19.571978.

5. Liu, L., Cui, Y., Chang, YZ. et al. Ferroptosis-related factors in the substantia nigra are associated with Parkinson’s disease. Sci Rep 13, 15365 (2023).

6. Buneeva O, Fedchenko V, Kopylov A, Medvedev A. Mitochondrial Dysfunction in Parkinson’s Disease: Focus on Mitochondrial DNA. Biomedicines. 2020 Dec 10;8(12):591.

7. Arotcarena ML, Bourdenx M, Dutheil N, Thiolat ML, Doudnikoff E, Dovero S, Ballabio A, Fernagut PO, Meissner WG, Bezard E, Dehay B. Transcription factor EB overexpression prevents neurodegeneration in experimental synucleinopathies. JCI Insight. 2019 Aug 22;4(16):e129719.

8. De Graeve F, Jagla T, Daponte JP, Rickert C, Dastugue B, Urban J, Jagla K. The ladybird homeobox genes are essential for the specification of a subpopulation of neural cells. Dev Biol. 2004 Jun 1;270(1):122–34.

9. Haque A, Polcyn R, Matzelle D, Banik NL. New Insights into the Role of Neuron-Specific Enolase in Neuro-Inflammation, Neurodegeneration, and Neuroprotection. Brain Sci. 2018 Feb 18;8(2):33.

10. Wang ZX, Li YL, Pu JL, Zhang BR. DNA Damage-Mediated Neurotoxicity in Parkinson’s Disease. Int J Mol Sci. 2023 Mar 28;24(7):6313.

11. Ross JM, Öberg J, Brené S, Coppotelli G, Terzioglu M, Pernold K, Goiny M, Sitnikov R, Kehr J, Trifunovic A, Larsson NG, Hoffer BJ, Olson L. High brain lactate is a hallmark of aging and caused by a shift in the lactate dehydrogenase A/B ratio. Proc Natl Acad Sci U S A. 2010 Nov 16;107(46):20087–92.

12. Yasuhara T, Shingo T, Kobayashi K, Takeuchi A, Yano A, Muraoka K, Matsui T, Miyoshi Y, Hamada H, Date I. Neuroprotective effects of vascular endothelial growth factor (VEGF) upon dopaminergic neurons in a rat model of Parkinson’s disease. Eur J Neurosci. 2004 Mar;19(6):1494–504.

13. Liu K, Li F, Han H, Chen Y, Mao Z, Luo J, Zhao Y, Zheng B, Gu W, Zhao W. Parkin Regulates the Activity of Pyruvate Kinase M2. J Biol Chem. 2016 May 6;291(19):10307–17.

14. Sadoul R. Bcl-2 family members in the development and degenerative pathologies of the nervous system. Cell Death Differ. 1998 Oct;5(10):805–15.

15. Quan Y, Wang J, Wang S, Zhao J. Association of the Plasma Long Non-coding RNA MEG3 With Parkinson’s Disease. Front Neurol. 2020 Nov 26;11:532891.

16. Li J, Chen L, Qin Q, Wang D, Zhao J, Gao H, Yuan X, Zhang J, Zou Y, Mao Z, Xiong Y, Min Z, Yan M, Wang CY, Xue Z. Upregulated hexokinase 2 expression induces the apoptosis of dopaminergic neurons by promoting lactate production in Parkinson’s disease. Neurobiol Dis. 2022 Feb;163:105605.

17. Li C, Ou R, Chen Y, Gu X, Wei Q, Cao B, Zhang L, Hou Y, Liu K, Chen X, Song W, Zhao B, Wu Y, Shang H. Mutation analysis of seven SLC family transporters for early-onset Parkinson’s disease in Chinese population. Neurobiol Aging. 2021 Jul;103:152.e1–152.e6.

18. Zhang HY, Jiang YC, Li JR, Yan JN, Wang XJ, Shen JB, Ke KF, Gu XS. Neuroprotective effects of insulin-like growth factor-2 in 6-hydroxydopamine-induced cellular and mouse models of Parkinson’s disease. Neural Regen Res. 2023 May;18(5):1099–1106.

19. Kannarkat GT, Boss JM, Tansey MG. The role of innate and adaptive immunity in Parkinson’s disease. J Parkinsons Dis. 2013;3(4):493–514.

20. Cathiard L, Fraulob V, Lam DD, Torres M, Winkelmann J, Krężel W. Investigation of dopaminergic signalling in Meis homeobox 1 (Meis1) deficient mice as an animal model of restless legs syndrome. J Sleep Res. 2021 Oct;30(5):e13311.

21. Liu J, Liu H, Zhao Z, Wang J, Guo D, Liu Y. Regulation of Actg1 and Gsta2 is possible mechanism by which capsaicin alleviates apoptosis in cell model of 6-OHDA-induced Parkinson’s disease. Biosci Rep. 2020 Jun 26;40(6):BSR20191796.

22. Gagliardi D, Pagliari E, Meneri M, Melzi V, Rizzo F, Comi GP, Corti S, Taiana M, Nizzardo M. Stathmins and Motor Neuron Diseases: Pathophysiology and Therapeutic Targets. Biomedicines. 2022 Mar 19;10(3):711.

23. Di Sano F, Piacentini M. Reticulon Protein-1C: A New Hope in the Treatment of Different Neuronal Diseases. Int J Cell Biol. 2012;2012:651805.

24. Geng X, Li S, Li J, Qi R, Zhong L, Yu H. MDMA targets miR-124/MEKK3 via MALAT1 to promote Parkinson’s disease progression. Mol Biol Rep. 2023 Nov;50(11):8889–8899.

25. Kokotos AC, Antoniazzi AM, Unda SR, Ko MS, Park D, Eliezer D, Kaplitt MG, Camilli P, Ryan TA. Phosphoglycerate kinase is a central leverage point in Parkinson’s Disease driven neuronal metabolic deficits. bioRxiv [Preprint]. 2023 Oct 10:2023.10.10.561760.

26. Li X, Cui XX, Chen YJ, Wu TT, Xu H, Yin H, Wu YC. Therapeutic Potential of a Prolyl Hydroxylase Inhibitor FG-4592 for Parkinson’s Diseases in Vitro and in Vivo: Regulation of Redox Biology and Mitochondrial Function. Front Aging Neurosci. 2018 Apr 27;10:121.

